# Good recovery of immunization stress-related responses presenting as cluster of stroke-like events following CoronaVac and ChAdOx1 vaccinations

**DOI:** 10.1101/2022.03.15.22272434

**Authors:** Metha Apiwattanakul, Narupat Suanprasert, Arada Rojana-Udomsart, Thanes Termglinchan, Chaichana Sinthuwong, Tasanee Tantirittisak, Suchat Hanchaiphiboolkul, Pantep Angchaisuksiri, Suphot Srimahachota, Jurai Wongsawat, Somjit Stiudomkajorn, Sasisopin Kiertiburanakul, Chonnamet Techasaensiri, Wannada Laisuan, Weerawat Manosuthi, Pawinee Doungngern, Wereyarmarst Jaroenkunathum, Teeranart Jivapaisarnpong, Apinya Panjangampatthan, Jirapa Chimmanee, Kulkanya Chokephaibulkit

## Abstract

**Background:** Immunization stress-related responses presenting as stroke-like symptoms may develop following COVID-19 vaccination. This study aimed to describe the clinical characteristics of immunization stress-related responses causing stroke-like events following COVID-19 vaccination.

**Methods:** We conducted a retrospective study of the secondary data of reported adverse events following COVID-19 immunization that presented with neurologic manifestations. Between March 1 and July 31, 2021, we collected and analyzed the medical records of 221 patients diagnosed with stroke-like symptoms following immunization. Demographic and medical data included sex, age, vaccine type, sequence dose, time to event, laboratory data, and recovery status as defined by the modified Rankin score (i.e., defining the degree of severity/dependence, with higher scores indicating greater disability). The affected side was evaluated for associations with the injection site.

**Results:** In total, 221 patients were diagnosed with immunization stress-related responses (stroke-like symptoms) following CoronaVac (Sinovac) or ChAdOx1 (AstraZeneca) vaccinations. Most patients (83.7%) were women. The median (interquartile range) age of onset was 34 (28–42) years in patients receiving CoronaVac and 46 (33.5–60) years in those receiving ChAdOx1. The median interval between vaccination and symptom onset for each vaccine type was 60 (16–960) min and 30 (8.8–750) min, respectively. Sensory symptoms were the most common symptomology. Most patients (53.8%) developed symptoms on the left side of the body; 99.5% of the patients receiving CoronaVac and 90% of those receiving ChAdOx1 recovered well (modified Rankin scores ≤2, indicating slight or no disability).

**Conclusions:** Immunization stress-related responses presenting as stroke-like symptoms can develop following COVID-19 vaccination. Symptoms that are more likely to occur on the injection side are transient (i.e., without permanent pathological deficits). Public education and preparedness are important for administering successful COVID-19 vaccination programs.

## Introduction

Immunization stress-related responses (ISRR) are defined by the World Health Organization (WHO) as symptoms and signs of bodily responses to vaccination (1). These events are not caused by side effects due to vaccine components but instead arise from the vaccination process/cascade.

Since the initiation of the COVID-19 vaccination campaign in Thailand on February 28, 2021, a total of 17,685,974 doses had been administered through the study follow-up date of July 31, 2021. Serious or concerning adverse events occurring within 30 days following vaccination as well as event clusters were reported to the Adverse Event Following Immunization (AEFI) committee at the Department of Disease Control (Thailand Ministry of Public Health). All cases were reviewed by the AEFI committee. Causality and relationships to vaccination were determined by consensus.

The first ISRR cluster was identified at the start of April 2021; the committee identified five patients with stroke-like symptoms. These patients were healthcare workers (HCWs; i.e., the first group to receive the vaccine in the country). Within one week, we identified many clusters of stroke-like symptoms reported following vaccination, all of which occurred in HCWs.

This information spread rapidly through social networks and caused vaccine hesitancy. Impurity or high vaccine specificity were suspected as probable causes. However, the relevant vaccine types were investigated and no problems were found. The AEFI committee convened to review these cases in a timely fashion and found that almost all of the reported cases were those of ISRR. Thus, herein, we report on the clinical features of clustered ISRR presenting with neurologic manifestations, with the goal of informing public health education, epidemic preparedness, and vaccine administration programs.

## Materials and methods

### Study design and participants

Thailand initiated a nationwide COVID-19 vaccination campaign on February 28, 2021. Medical records were systematically reviewed following the report of each AEFI. In some cases of doubt, the treating clinician was contacted for additional information. The AEFI committee convened in a timely manner to determine causality as well as the relationships of the presenting symptomology with vaccination status and timing.

We conducted a retrospective study within secondary data received from the national AEFI surveillance program (AEFI-DDC). These data were reviewed by the AEFI committee. Specifically, we evaluated ISRR with a neurologic presentation that occurred between March 1 and July 31, 2021. The data evaluated in this secondary analysis were obtained without direct contact with patients or their attending healthcare workers. Neurological ISRR were diagnosed based on the presence of clinical neurological symptoms compatible with dissociative neurological symptom reactions (1).

The data used for this analysis, including sex, age, vaccine type, sequence dose, time to event, clinical manifestations, laboratory data, and recovery status (which was defined according to the patients’ modified Rankin scale [mRS] scores, with higher scores indicating a greater level of disability) were abstracted from the study database provided by the AEFI committee. The mRs scores were selected as the primary outcome measure with respect to defining recovery status. A good outcome was defined as an mRS score of 0–2 (indicating slight or no disability), a poor outcome was defined as an mRS score of 3–5 (indicating moderate to severe disability), and mortality was categorized as an mRS score of 6.

This study was approved by the Ethical Review Committee for Research in Human Subjects at the Thailand Ministry of Public Health approval No. 15/2564. The requirement for informed consent was waived due to the retrospective nature of the current study and the fact that we conducted a secondary analysis of anonymized and deidentified data.

### Statistical analysis

When evaluating baseline demographic and medical data, we report medians and interquartile ranges (IQR) for continuous data (some variables were not normally distributed), whereas categorical variables are presented as counts and percentages. Demographic and medical data were dichotomized by vaccine type (CoronaVac [Sinovac Biotech, Beijing, China], ChAdOx1 [AstraZeneca, Cambridge, UK]). Proportions of categorical variables were compared across vaccine groups using the X^2^ test. Mean body mass index (BMI) was compared with the theoretical mean (BMI = 25 kg/m^2^) using a one-sample t-test. We also evaluated the distributions and characteristics of adverse events by injection site. All statistical analyses were performed using GraphPad Prism 9 software (San Diego, CA, USA). The threshold for statistical significance was set at a two-sided *p* value of less than 0.05.

## Results

Following a total of 17,685,974 COVID-19 vaccine doses that were administered as of July 31, 2021 (including 8,699,803 doses of the CoronaVac vaccine and 8,000,079 doses of the ChAdOx1 vaccine), 293 patients with serious neurological complications were reported to the AEFI committee. These included 278 and 15 patients who had received the CoronaVac and ChAdOX1 vaccines, respectively.

ISRR was diagnosed in 263 patients. Other neurological diseases were identified in 18 patients, including five patients with stroke (two with ischemic stroke and three with hemorrhagic stroke), four patients with neuropathy, four with provoked seizure, one with severe headache, one with severe myalgia, one with facial edema and numbness, one experiencing syncope, and one with aggravated back pain due to spinal stenosis. For 12 patients who reported severe neurological symptoms, the AEFI committee determined the causality as inconclusive due to incomplete medical records. Of the 263 patients reported to have ISRR, 221 had adequate data available for this secondary analysis. All patients had normal findings on brain CT computed tomography (CT) and/or magnetic resonance imaging (MRI) scans. Table 1 shows demographic data, clinical symptomology, and health outcomes in the identified patients experiencing adverse vaccine-associated events.

**Table 1.**
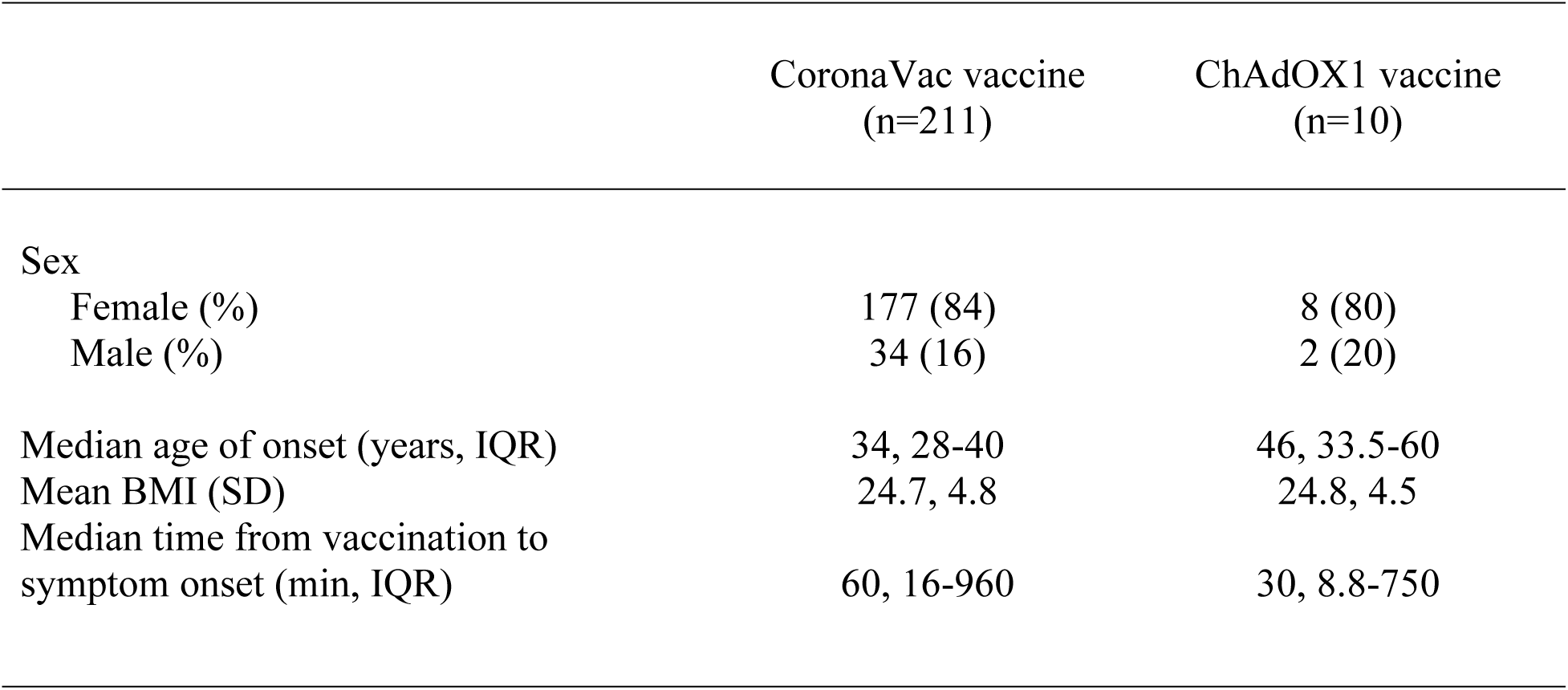
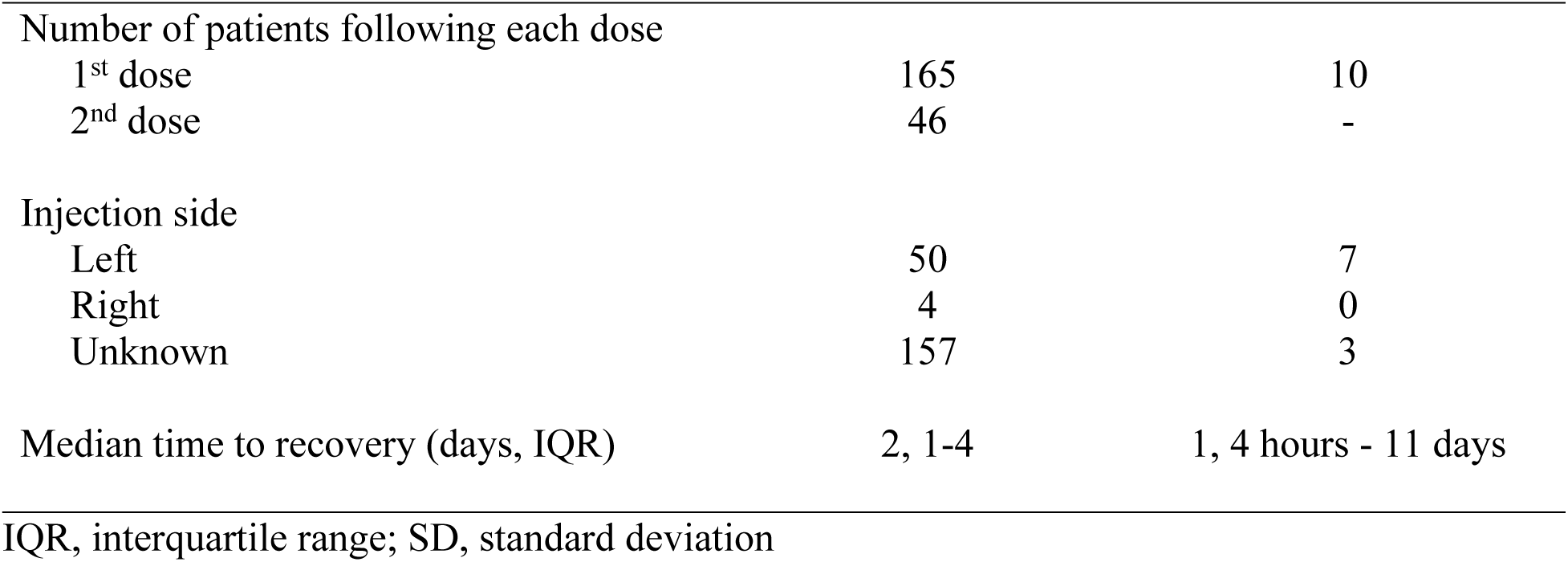
Clinical and demographic data for immunization stress-related responses (ISRR)

Most of these patients were females (185/221, 87%). The median age of onset was 34 years (IQR, 28–42) for patients who received the CoronaVac vaccine and 46 years (33.5–60) for those who received the ChAdOx1 vaccine. The median interval between vaccination and symptom onset was 60 min (IQR, 16–960) for the CoronaVac vaccine and 30 min (8.8–750) for the ChAdOX1 vaccine. Most of these adverse events (175/221, 79.2%) occurred following the first vaccination dose. The median time from injection to recovery was 2 days (IQR, 1–4) and 1 day (IQR, 4 h to 11 days) for CoronaVac and ChAdOX1 recipients, respectively.

A total of 209 patients (94.5%) had sensory, motor, or combined sensorimotor symptoms. Of these, 144 (68.9%) patients developed symptoms on the left side of the body (injection side). Of the 50 patients whose injection side was known, 45 (90%) developed symptoms on the same side as the injection site. Table 2 showed clinical characteristic with respect to neurological symptoms stratified by the vaccine injection site. In CoronaVac recipients (211 patients), 74.8% were fully recovered and 99.5% had good outcomes (mRS 0–2). Only one patient had an mRS score of 3. This patient had clinical symptoms and laboratory data compatible with ISRR; the causes of this adverse event are currently being explored by a multidisciplinary team. All 10 ChAdOX1 recipients had good outcomes (mRS 0–2). The mean BMI of participants with information on this variable (214 patients) was 24.67 kg/m^2^ (standard deviation, 4.78). There was no association between ISRR and overweight/obesity (BMI ≥25 kg/m^2^; p=0.31, 95% confidence interval [CI] −0.9750 to 0.3150).

**Table 2.** Clinical characteristic with respect to neurological symptoms stratified by the vaccine injection site.

|  | Left side injection<br>(n=57) | Right side injection<br>(n=4) | Unknown<br>(n=160) |
| --- | --- | --- | --- |
| Lateralization of symptoms |  |  |  |
| Left side | 42 | 1 | 101 |
| Right side | 4 | 3 | 29 |
| Bilateral | 6 | - | 14 |
| Unknown | 2 | - | 7 |
| Non-stroke-like | 3 | - | 9 |
| Sensory symptoms |  |  |  |
| Sensory (NA) | 2 | - | 7 |
| Sensory, left side | 20 | 1 | 59 |
| Sensory, right side | 1 | 2 | 19 |
| Sensory, bilateral | 2 | - | 5 |
| Motor symptoms |  |  |  |
| Motor (NA) | - | - | - |
| Motor, left | 2 | - | 5 |
| Motor, right | 1 | 1 | 2 |
| Motor, bilateral | 1 | - | 5 |
| Sensorimotor (SM) symptoms |  |  |  |
| SM (NA) | - | - | - |
| SM, left | 20 | - | 37 |
| SM, right | 2 | - | 8 |
| SM, bilateral | 3 | - | 4 |

NA, not applicable

## Discussion

There are several well-recognized neurological complications associated with many forms of vaccination, including Guillain-Barré syndrome and acute disseminated encephalomyelitis. These effects can be explained by the immunological mimicry of vaccine antigens to the relevant myelin protein. However, stroke has not been recognized as an AEFI. Moreover, to our knowledge, stroke-like syndromes occurring as clustered events have not been reported as adverse vaccine-associated events to date.

ISRR is considered an alternative term for conversion disorder and/or dissociative neurological symptomology. Functional neurological disorders (FND) were described thoroughly in a recent report (2). FND are not malingering and are rather neurological deficits that are not caused by structural lesions and are instead characterized by changes in the functionality of neurons and glia as well as physiologic changes occurring in specific brain regions. Symptoms and signs in these patients are real and nonvolitional (in contrast to what has been perceived in the past). These may be the typical responses of the body to the normal physiological processes occurring after immunization (i.e., pain or inflammation), and stress or fear of side effects may aggravate this symptomology. Well-characterized functional disorders in other systems include irritable bowel syndrome, vasovagal syncope, stress-associated dyspepsia, and stress-aggravated migraine headache (1).

A theory that may explain the occurrence of vaccine-associated stroke-like symptomology based on a mechanism involving complex regional pain syndrome and FND has been proposed in a prior report (3). We note that most patients develop clinical symptoms of dysesthesia or numbness in the same limb as the immunization site. We also note that the patients with stroke-like symptoms reported in our cohort had good prognoses; most patients recovered fully and none of them had structural deficits.

Based on our findings, we encourage more comprehensive health education regarding this ISRR for both vaccine recipients and administering healthcare workers, who are responsible for effective clinical decision-making, meticulously monitoring patients’ health, and reassuring patients that their symptoms are normal responses and that they will recover. These adverse events should not impede any vaccination campaigns, especially during the pandemic. Appropriate follow-up investigation is also essential so as not to miss real structural neurological deficits as well as to provide appropriate investigation and treatment when necessary.

The ISRR clusters evaluated in our study were reported at the time the worldwide vaccine phobia wave initially emerged due to widespread and frequently inaccurate information regarding the side effects of vaccination, which was highly prevalent on social media at the time. In Thailand, case clustering has been reported throughout the country. Vaccine constituents were the suspected culprits at the start of the vaccine phobia wave. Moreover, many patients were diagnosed with stroke and received unnecessary thrombolytic or antiplatelet therapy despite normal findings on brain CT and/or MRI. These and similar effects and phenomena severely impede the efficacy and reach of vaccination campaigns.

Following appropriate investigation and a confirmation of the occurrence of ISRR, preparedness and management strategies were introduced on a nationwide scale. The number of reported cases was thus reduced substantially. WHO consultation and preparedness guidance were also extremely helpful in facilitating this process.

The first case report of stroke-like symptomology occurring after CoronaVac vaccination in this country described an acute prolonged motor aura (4). The patient in this case developed visual symptoms within 15 min after vaccination, followed by left arm numbness and weakness. CT and MRI findings were normal. We performed single-photon emission computerized tomography with Tc-99 ethyl cysteinate dimer, demonstrating relative hypoperfusion to the right cerebral hemisphere. However, in our study, most patients did not develop headaches. We also note that adverse events were likely to be lateralized to the side of injection. We propose that the underlying mechanisms for vaccine-associated ISSR may be of peripheral origin (e.g., pain following vaccination) and may also be integrated with an autonomic response (i.e., a response resembling a reflex sympathetic response, as in complex regional pain syndrome). According to previous research, this symptomology may also involve the central nervous system, as demonstrated by abnormalities in the postcentral gyrus and inferior parietal cortex, which are responsible for afferent information processing (5, 6). This may explain the transient clinical weakness seen in some of our enrolled patients.

ISRR was initially perceived as psychogenic within the medical community. However, these symptoms are real and not iatrogenic. The initial confusion arose from the presenting symptomology being caused by functional physiologic changes rather than by structural damage. The same situation has been reported with many vaccines in the past (7-11). Many underlying mechanisms have been proposed, including pain due to the vaccination process and/or inflammation occurring after vaccination, which may activate the peripheral nerves and/or the sympathetic nervous system, as explained in a prior description of the proposed complex regional pain syndrome mechanism (3).

Although we did not have information regarding the injection site in many cases, most injections are likely performed in the non-dominant arm. This is consistent with the majority of reported symptoms occurring on the left side of the body. Conversion disorder and/or psychogenic weakness have been suspected in the past. However, evidence based on functional imaging of the brain was demonstrated in these aforementioned cases (5).

Recently, two cases were reported wherein the patients developed a clinical level of stroke-like weakness after receiving an mRNA-based COVID-19 vaccine (12). Another case report described hemiparesis on the same side as the injection site that lasted for 40 min. However, this patient developed hypoesthesia on the alternate side and the neurological examination revealed midline splitting of the sensory deficit. This patient also had normal brain CT and MRI findings. The authors hypothesized that increased attention toward body signals and abnormal expectations regarding the symptoms of vaccination-induced injury may be responsible for this symptomology (13). This report confirms that the process of incident vaccine-associated neurological disorders is more likely due to the vaccination process rather than to specific vaccine constituents, since these events have occurred within various vaccine platforms.

As COVID-19 mass vaccination has been widely implemented among high-risk populations with underlying diseases, we found that some true cases of stroke were simply temporally associated with vaccination within the current study. This caveat provides precautionary information and informs the provision of appropriate timely treatment (vs. empirically treating ISRR in the absence of differential diagnosis).

We acknowledge several limitations of this report. Specifically, this evaluation was based on a retrospective study within the secondary data and we may have therefore missed some critical information (especially with respect to outcome data). The reported data regarding female predominance could be due to the initial target population of the nationwide vaccine campaign, which initially comprised HCWs who received preferential vaccination at the start of the vaccine campaign, most of whom were young females (e.g., nurses, therapists). Moreover, we were unable to identify the factors associated with ISRR in this descriptive study and instead present a descriptive evaluation only. Future studies should evaluate risk factors and causality more comprehensively using multivariate modeling and a prospective design.

## Conclusions

To our knowledge, our study reports the largest series regarding ISRR associated with neurological symptoms identified to date. Stroke-like symptoms are more likely to occur on the same side as the injection site, which may be explained by the triggering of painful stimuli and exaggerated self-attention as the main suspected mechanisms mediating the development of this transient symptomology. A high degree of awareness with respect to this symptomology is important to avoid over-investigation, which could result in unneeded costs and complications. It is important to provide effective and comprehensive education regarding these events to the public as well as to the administering HCWs. These occurrences are real but transient and almost invariably present with excellent recovery prospects. This information as well as more comprehensive information obtained in future studies will undoubtedly reduce vaccine hesitancy and will help ensure a successful nationwide vaccination program as well as informing interventions on a global scale.

## Data Availability

All relevant data are within the manuscript and its Supporting Information files.

## Author Contributions

Dr. Apiwattanakul had full access to all the data in the study and takes responsibility for the integrity of the data and the accuracy of the data analysis.

Concept and design: Dr. Suanprasert, Dr. Rojana-Udomsart, Dr. Termglinchan, Dr. Sinthuwong, Dr. Tantirittisak, Dr. Hanchaiphiboolkul

Acquisition, analysis, or interpretation of data: All authors.

Critical revision of manuscript for important intellectual content: Dr. Hanchaiphiboolkul, Dr. Chokephaibulkit

Statistical analysis: Dr. Apiwattanakul, Dr. Hanchaiphiboolkul

Administrative, technical, or material support: Dr. Doungngern, Ms. Panjangampatthan, Ms. Chimmanee

Supervision: Dr. Tantirittisak, Dr. Hanchaiphiboolkul, Dr. Chokephaibulkit

